# Effectiveness of the COVID-19 vaccines against severe disease with Omicron sub-lineages BA.4 and BA.5 in England

**DOI:** 10.1101/2022.08.31.22279444

**Authors:** Freja C. M. Kirsebom, Nick Andrews, Julia Stowe, Mary Ramsay, Jamie Lopez Bernal

## Abstract

The Omicron sub-lineages BA.4 and BA.5 were first detected in England in April 2022. A case surge followed despite England having recently experienced waves with BA.1 and BA.2. This study used a whole population test-negative case-control study design to estimate the effectiveness of the vaccines currently in use as part of the UK COVID-19 vaccination programme against hospitalisation following infection with BA.4 and BA.5 as compared to BA.2 during a period of co-circulation. Incremental VE was estimated in those vaccinated with either a third or fourth dose as compared to individuals with waned immunity who had received their second dose at least 25 weeks prior. Vaccination status was included as an independent variable and effectiveness was defined as 1-odds of vaccination in cases/odds of vaccination in controls. During the study period, there were 32,845 eligible tests from hospitalised individuals. Of these, 25,862 were negative (controls), 3,432 were BA.2, 273 were BA.4, 947 were BA.5 and 2,331 were either BA.4 or BA.5 cases. There was no evidence of reduced VE against hospitalisation for BA.4 or BA.5 as compared to BA.2. The incremental VE was 56.8% (95% C.I.; 24.0-75.4%), 59.9% (95% C.I.; 45.6-70.5%) and 52.4% (95% C.I.; 43.2-60.1%) for BA.4, BA.5 and BA.2, respectively, at 2 to 14 weeks after a third or fourth dose. VE against hospitalisation with BA.4/5 or BA.2 was slightly higher for the mRNA-1273 booster than the BNT162b2 booster at all time-points investigated, but confidence intervals overlapped. These data provide reassuring evidence of the protection conferred by the current vaccines against severe disease with BA.4 and BA.5.

---

The Omicron sub-lineages BA.4 and BA.5, identified in South Africa in early 2022 [1], were first detected in England in April 2022 [2]. A case surge followed despite England having recently experienced waves with BA.1 and BA.2. BA.4 and BA.5 have identical spike proteins most similar to that of BA.2 but with additional mutations including the 69–70 deletion, L452R, F486V and wild-type amino acid at position Q493 [1]. Neutralisation assays have found BA.4 and BA.5 display increased evasion of antibodies from plasma of vaccinated or BA.1 infected individuals, as compared to BA.2 [3-5]. Recent data from Denmark and Portugal have found that the odds of being vaccinated did not differ amongst BA.5 and BA.2 cases [6, 7]. The Portuguese study did find lower VE against hospitalisation for BA.5 using a cohort study design [6].

The UK COVID-19 vaccination program has been in place since December 2020 with primary courses of two doses of BNT162b2 (Pfizer-BioNTech), ChAdOx1-S (AstraZeneca) or mRNA-1273 (Moderna). Third doses with either BNT162b2 or a half dose (50μg) of mRNA-1273 were offered to all adults by December 2021. Fourth doses were offered to those at risk and those aged 75 years and older from March 2022.

We used a test-negative case-control (TNCC) study design to investigate VE against hospitalisation for BA.4. BA.5 and BA.2 during a period of co-circulation, as previously described [8-10]. Vaccination status was included as an independent variable and effectiveness was defined as 1-odds of vaccination in cases/odds of vaccination in controls. For BA.2, VE following a third or fourth dose was comparable (Supplementary Table 2). Therefore, incremental VE was estimated in those vaccinated with either a third or fourth dose as compared to individuals with waned immunity who had received their second dose at least 25 weeks prior (full details in Supplementary Appendix).

Between 18 April and 17 July 2022 there were 32,845 eligible tests from individuals hospitalised for at least 2 days and with a respiratory code in the primary diagnosis field. Of these, 25,862 were negative (controls), 3,432 were BA.2, 273 were BA.4, 947 were BA.5 and 2,331 were either BA.4 or BA.5 cases (Supplementary Table 3).

There was no evidence of reduced VE against hospitalisation for BA.4 or BA.5 as compared to BA.2 (Figure 1, Supplementary Table 4&5). In those who had received their third or fourth dose 2 to 14 weeks ago, the incremental VE as compared to those who were 25 or more weeks post their second dose was 56.8% (95% C.I.; 24.0-75.4%) and 59.9% (95% C.I.; 45.6-70.5%) for BA.4 and BA.5, respectively, and 52.4% (95% C.I.; 43.2-60.1%) for BA.2 (Supplementary Table 4). Incremental VE waned to 1.5% (95% C.I.; -63.1-40.5%), 23.3% (95% C.I.; -1.5-42.1%) and 9.8% (95% C.I.; -6.0-23.3%) for BA.4, BA.5 and BA.2 at 25 or more weeks.

**Figure 1.**
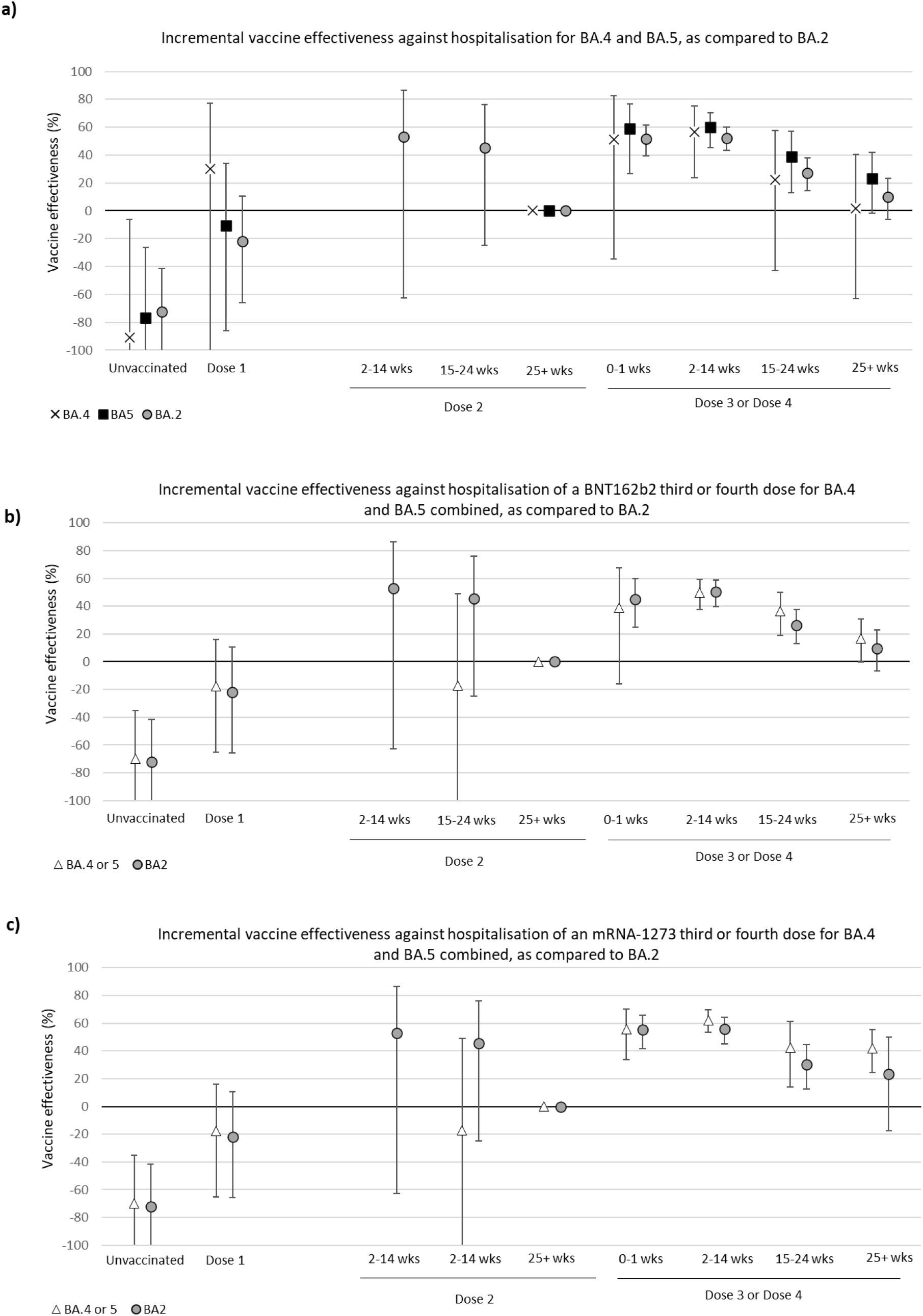
Incremental VE against hospitalisation with BA.4, BA.5 and BA.2 in England, compared using individuals who received a second dose at least 25 weeks prior as the baseline. VE for all vaccine manufacturers combined (a). VE of a BNT162b2 (b) or mRNA-1273 (c) third or fourth dose.

To investigate VE by manufacturer, we stratified by final (third or fourth) dose manufacturer (BNT162b2 or mRNA-1273). BA.4 and BA.5 cases were combined for precision. There was no difference in VE against hospitalisation for BA.4/5 as compared to BA.2 (Figure 1b&c, Supplementary Table 5). VE against hospitalisation with BA.4/5 or BA.2 was slightly higher for mRNA-1273 than BNT162b2 at all time-points investigated, but confidence intervals overlapped. Incremental VE against hospitalisation with BA.4/5 was 62.2% (95% C.I.; 53.2-69.4%) and 49.6% (95% C.I.; 37.5-59.4%) for mRNA-1273 and BNT162b2, respectively, at 2 to 14 weeks after receiving a third or fourth dose (Figure 1b&c, Supplementary Table 5). This decreased to a VE of 16.7% (95% C.I.; -0.3-30.7%) and 42.0% (95% C.I.; 24.5-55.5%) for BNT162b2 and mRNA-1273, respectively, at 15 to 24 weeks.

These data provide reassuring evidence of the protection conferred by the current vaccines against severe disease with BA.4 and BA.5; we found no difference in VE as compared to BA.2 and BNT162b2 and mRNA-1273 boosters provided similarly high levels of protection. This contradicts pre-printed data from a cohort study in Portugal which found VE against severe outcomes was lower for BA.5 [6]. This may be due to the small size of the Portuguese study, methodological differences, or differences in classifying hospitalised cases. Here, we use a strict definition as we have previously found broader definitions give lower estimates which likely reflect VE against symptomatic disease [9]. Risk factor status and previous infection will also impact VE; these analyses include adjustment by most recent previous variant and by the risk factor groups offered early vaccination which the Portuguese study did not. Differences in testing policies between countries will also impact local ability to adjust for factors such as previous infection.

## Data Availability

All data produced in the present work are contained in the manuscript. Data cannot be made publicly available for ethical and legal reasons, i.e. public availability would compromise patient confidentiality as data tables list single counts of individuals rather than aggregated data

## Authors’ contributions

FCMK and JLB wrote the manuscript. JLB, NA and MR conceptualised the study. FCMK and JS curated the data. FCMK and NA conducted the formal analysis. All co-authors reviewed the manuscript.

## Ethics committee approval

UKHSA has legal permission, provided by Regulation 3 of The Health Service (Control of Patient Information) Regulations 2002, to process patient confidential information for national surveillance of communicable diseases and as such, individual patient consent is not required to access records.

