## Supplementary Appendix for "Effectiveness of the COVID-19 vaccines against severe disease with Omicron sub-lineages BA.4 and BA.5 in England"

### Methods

#### Study Design

A test negative case control design was used to estimate vaccine effectiveness against hospitalisation with Omicron sub-lineages BA.2, BA.4 and BA.5 in individuals aged 18 years and older. The odds of vaccination in Pillar 1 PCR positive cases were compared to the odds of vaccination in individuals who tested negative in England.

### Data Sources

#### COVID-19 Testing Data

Prior to the 01 April 2022, PCR testing for SARS CoV-2 in England was undertaken by hospital and public health laboratories (Pillar 1), as well as by community testing (Pillar 2). Pillar 1 testing is PCR testing in public health laboratories and NHS hospitals and was available for inpatients and others presenting to secondary care as well as health and care workers. Pillar 2 testing included lateral flow tests (LFTs) and PCR tests. In Pillar 2, LFTs were available to everyone, and PCR testing was available to anyone with symptoms consistent with COVID-19 (high temperature, new continuous cough, or loss or change in sense of smell or taste), anyone who was a contact of a confirmed case, care home staff and residents, and to those who self-tested as positive using an LFT. Since 01 April 2022, community testing has been scaled down and PCR tests are no longer freely available for most people. LFT tests are available for people with certain health conditions, for those going into hospital and for those who work in the NHS or in adult social care. Since 01 April 2022, PCR testing is still being undertaken via Pillar 1.

Individuals who were PCR tested in Pillar 1 between 18 April 2022 and 17 July 2022 (a period of co-circulation of BA.2, BA.4, and BA.5) were included in the final analysis. Any negative tests taken within 7 days of a previous negative test were excluded as these likely represent the same episode. Negative tests taken within 21 days of any subsequent positive test (LFT or PCR, Pillar 1 or Pillar 2) were also excluded as chances are high that these are false negatives. Positive and negative tests within 90 days of any previous positive test (LFT or PCR, Pillar 1 or Pillar 2) were also excluded. After linkage to the admission hospital data negatives tests within 21 days of a previous negative were also excluded, and, for individuals who had more than one linked negative test, one was selected at random in the study period. The test date of the most recent previous positive test (Pillar 1 or Pillar 2), excluding tests within 90 days, was extracted for all individuals to allow adjustment for prior infection.

#### Vaccination Data

The National Immunization Management System (NIMS) contains demographic information on the whole population of England who are registered with a general practice physician in England and is used to record all COVID-19 vaccinations. NIMS was accessed for dates of vaccination and manufacturer, sex, date of birth, ethnicity, and residential address. Addresses were used to determine index of multiple deprivation quintile and were also linked to Care Quality Commission registered care homes using the unique property reference number. Data on geography (NHS region), risk group status, clinically extremely vulnerable status, and health/social care worker were also extracted from the NIMS. Clinical risk groups included a range of chronic conditions as described in the Green Book [1], whereas the clinically extremely vulnerable group included persons who were considered to be at the highest risk for severe COVID-19, including those with immunosuppressed conditions and those with severe respiratory disease. Third doses were given at least 84 days after a second dose and administered after 13 September 2021. Individuals with a heterologous primary schedule or fewer than 19 days between their first and second dose were excluded. Fourth doses given from 01 January 2022 and at least 84 days after a third dose were counted, with those given the dose with shorter intervals or prior to 01 January 2022 excluded.

Testing data were linked to NIMS on 01 August 2022 using combinations of the unique individual National Health Service (NHS) number, date of birth, surname, first name, and postcode using deterministic linkage.

### Identification of Omicron Variants and assignment to cases

Sequencing of PCR positive samples is undertaken through a network of laboratories, including the Wellcome Sanger Institute. Whole-genome sequences are assigned to UKHSA definitions of variants based on mutations [2]. Cases were defined as BA.2, BA.4 or BA.5 based on whole genome sequencing. From 18 April 2022 to 23 May 2022 the positive predictive value (PPV) of a sequenced test being BA.2 was at least 80% so not sequenced tests were classified as BA.2. From 20 June 2022 onwards, the PPV of a sequenced test being either BA.4 or BA.5 was at least 80%, so from then on tests not sequenced were classified as BA.4 or BA.5 and included in analyses which combined BA.4 and BA.5 cases. Where subsequent positive tests within 14 days included sequencing information, this information was used to classify the variant.

### Hospital Admission Data

#### Secondary Care Hospital Admission Data (SUS)

SUS is the national electronic database of hospital admissions that provides timely updates of ICD-10 codes for completed hospital stays for all NHS hospitals in England. Up to 24 ICD-10 diagnoses fields can be completed in SUS for each admission with the first diagnosis field indicating the primary reason for admission. Hospital inpatient admissions for a range of acute respiratory illnesses were identified from SUS and were linked to the testing data on 08 August 2022 using NHS number and date of birth as previously described [3]. Admissions with an ICD-10 coded ARI discharge diagnosis (Supplementary Table 1) in any diagnosis field were identified where the sample was taken 1 day before and up to 2 days after the admission. Length of stay was calculated as date of discharge – date of admission. Where multiple admissions linked to the same sample date, the first admission after the sample date was retained and episode length calculated by summing the stay length for each admission. Data were then restricted to those with ARI in the first diagnosis field and where the length of stay was at least two days. The data was restricted to tests up to 17 July 2022 to account for delays in the SUS data recording.

### Statistical Analysis

Logistic regression was used, with the PCR test result as the dependent variable and cases being those testing positive (stratified in separate analyses as either BA.2, BA.4, BA.5, or BA.4 and BA.5) and controls being those testing negative. Vaccination status at the date of test (or onset if available) was included as an independent variable and effectiveness defined as 1- odds of vaccination in cases/odds of vaccination in controls.

Vaccine effectiveness was adjusted in logistic regression models for age (ages 18-19, then 20 through to 89 in five-year bands, then everyone age 90 years or older), sex, index of multiple deprivation (quintile), ethnic group, care home residence, , geographic region (NHS region), period (week of test), health and social care worker status, clinical risk group status, clinically extremely vulnerable, and variant of most recent previous infection (none, wild-type, Alpha, Delta or Omicron

– variant status determined by date of test). These factors were all considered potential confounders so were included in all models.

Analysis combined all vaccine manufacturers (ChAdOx1, BNT162b2 or mRNA-1273 for one dose or two doses, and BNT162b2 or mRNA-1273 (half-dose) for third and fourth doses) or were stratified by the manufacturer of the third and fourth dose (whichever was the final dose). Heterologous primary schedules and ChAdOx1 primary course followed by ChAdOx1 third dose recipients were excluded.

Vaccine effectiveness was assessed in the unvaccinated, in dose 1 recipients, in intervals of 0 to 1 weeks, 2 to 14 weeks and 15 to 24 weeks post a second dose, and in intervals of 0 to 1 weeks, 2 to 14 weeks, 15-24 weeks and 25 or more weeks post a third or fourth dose. Vaccine effectiveness was estimated relative to those with waned immunity (25 or more weeks post their second dose).

Supplementary Table 1. SUS Acute respiratory illness ICD10 code list.

| SUS Acute respiratory illness ICD10 code list | |
|---|---|
| J04* | Acute laryngitis and tracheitis |
| J09* | Influenza due to identified avian influenza virus |
| J10* | Influenza with pneumonia, other influenza virus identified |
| J11* | Influenza with pneumonia, virus not identified |
| J12* | Viral pneumonia, not elsewhere classified |
| J13* | Pneumonia due to Streptococcus pneumoniae |
| J14* | Pneumonia due to Haemophilus influenzae |
| J15* | Bacterial pneumonia, not elsewhere classified |
| J16* | Pneumonia due to other infectious organisms, not elsewhere classified |
| J17* | Pneumonia in diseases classified elsewhere |
| J18* | Pneumonia, organism unspecified |
| J20* | Acute bronchitis |
| J21* | Acute bronchiolitis |
| J22* | Unspecified acute lower respiratory infection |
| J80* | ARDS (related to respiratory infection) |
| U07* | COVID-19, virus identified and not identified |
| U04* | Severe acute respiratory syndrome (SARS) |

Supplementary Table 2. Vaccine effectiveness estimates against hospitalisation with BA.2 for individuals aged 18 years and older in England. Third and fourth dose VE is estimated relative to a second dose at least 25 weeks prior.

| Doses | Interval (weeks) | Controls | Cases | VE % (95% CI) |
|---|---|---|---|---|
| Unvaccinated | | 1,235 | 349 | -72.0 (-109 to -41.5) |
| Dose 1 | >0 | 406 | 73 | -21.8 (-65.7 to 10.4) |
| Dose 2 | 0-1 | 4 | 1 | n too small |
| | 2-14 | 35 | 3 | n too small |
| | 15-24 | 78 | 8 | 45.5 (-24.4 to 76.1) |
| | 25+ | 1,645 | 258 | Baseline |
| Dose 3 | 0-1 | 54 | 3 | 61.9 (-27.8 to 88.6) |
| | 2-14 | 520 | 46 | 56.7 (38.6 to 69.4) |
| | 15-24 | 4,636 | 928 | 27.5 (14.7 to 38.4) |
| | 25+ | 8,004 | 1,079 | 9.6 (-6.4 to 23.1) |
| Dose 4 | 0-1 | 1,389 | 160 | 51.3 (38.8 to 61.2) |
| | 2-14 | 7,280 | 486 | 51.7 (42.2 to 59.7) |
| | 15+ | 576 | 38 | 18.4 (-20.5 to 44.7) |

Supplementary Table 3. Descriptive characteristics of eligible tests from hospitalised individuals.

| | | | Overall | | Negative | | BA.2 | | BA.4 | | BA.5 | | BA.4 or BA.5 | |
|---|---|---|---|---|---|---|---|---|---|---|---|---|---|---|
| | | | **n** | **%** | **n** | **%** | **n** | **%** | **n** | **%** | **n** | **%** | **n** | **%** |
| | **Test Result** | **Interval (weeks)** | **32,845** | **100.0%** | **25,862** | **78.7%** | **3,432** | **10.4%** | **273** | **0.8%** | **947** | **2.9%** | **2,331** | **7.1%** |
| **Vaccination Status and intervals after vaccine** | Unvaccinated | | 1,993 | **6.1%** | 1,235 | **4.8%** | 349 | **10.2%** | 33 | **12.1%** | 107 | **11.3%** | 269 | **11.5%** |
| | Dose 1* | >0 | 577 | **1.8%** | 406 | **1.6%** | 73 | **2.1%** | 4 | **1.5%** | 24 | **2.5%** | 70 | **3.0%** |
| | Dose 2* | 0-1 | 5 | **0.0%** | 4 | **0.0%** | 1 | **0.0%** | 0 | **0.0%** | 0 | **0.0%** | 0 | **0.0%** |
| | | 2-14 | 41 | **0.1%** | 35 | **0.1%** | 3 | **0.1%** | 1 | **0.4%** | 1 | **0.1%** | 1 | **0.0%** |
| | | 15-24 | 97 | **0.3%** | 78 | **0.3%** | 8 | **0.2%** | 2 | **0.7%** | 2 | **0.2%** | 7 | **0.3%** |
| | | 25+ | 2,186 | **6.7%** | 1,645 | **6.4%** | 258 | **7.5%** | 20 | **7.3%** | 75 | **7.9%** | 188 | **8.1%** |
| | Dose 3 or 4** | 0-1 | 525 | **1.6%** | 446 | **1.7%** | 64 | **1.9%** | 1 | **0.4%** | 3 | **0.3%** | 11 | **0.5%** |
| | BNT162b2 | 2-14 | 4,739 | **14.4%** | 3,959 | **15.3%** | 335 | **9.8%** | 29 | **10.6%** | 125 | **13.2%** | 291 | **12.5%** |
| | | 15-24 | 5,282 | **16.1%** | 4,223 | **16.3%** | 805 | **23.5%** | 24 | **8.8%** | 71 | **7.5%** | 159 | **6.8%** |
| | | 25+ | 9,746 | **29.7%** | 7,375 | **28.5%** | 1,052 | **30.7%** | 107 | **39.2%** | 360 | **38.0%** | 852 | **36.6%** |
| | Dose 3 or 4** | 0-1 | 1,140 | **3.5%** | 997 | **3.9%** | 99 | **2.9%** | 4 | **1.5%** | 13 | **1.4%** | 27 | **1.2%** |
| | mRNA-1273 | 2-14 | 4,508 | **13.7%** | 3,841 | **14.9%** | 197 | **5.7%** | 32 | **11.7%** | 125 | **13.2%** | 313 | **13.4%** |
| | | 15-24 | 1,199 | **3.7%** | 989 | **3.8%** | 161 | **4.7%** | 5 | **1.8%** | 14 | **1.5%** | 30 | **1.3%** |
| | | 25+ | 807 | **2.5%** | 629 | **2.4%** | 27 | **0.8%** | 11 | **4.0%** | 27 | **2.9%** | 113 | **4.8%** |
| **Age** | 18-19 | | 80 | **0.2%** | 61 | **0.2%** | 3 | **0.1%** | 1 | **0.4%** | 4 | **0.4%** | 11 | **0.5%** |
| | 20-24 | | 228 | **0.7%** | 142 | **0.5%** | 32 | **0.9%** | 4 | **1.5%** | 15 | **1.6%** | 35 | **1.5%** |
| | 25-29 | | 323 | **1.0%** | 195 | **0.8%** | 48 | **1.4%** | 9 | **3.3%** | 20 | **2.1%** | 51 | **2.2%** |
| | 30-34 | | 406 | **1.2%** | 289 | **1.1%** | 41 | **1.2%** | 13 | **4.8%** | 22 | **2.3%** | 41 | **1.8%** |
| | 35-39 | | 465 | **1.4%** | 353 | **1.4%** | 51 | **1.5%** | 7 | **2.6%** | 14 | **1.5%** | 40 | **1.7%** |
| | 40-44 | | 475 | **1.4%** | 369 | **1.4%** | 35 | **1.0%** | 5 | **1.8%** | 23 | **2.4%** | 43 | **1.8%** |
| | 45-49 | | 683 | **2.1%** | 527 | **2.0%** | 71 | **2.1%** | 3 | **1.1%** | 20 | **2.1%** | 62 | **2.7%** |
| | 50-54 | | 1,068 | **3.3%** | 855 | **3.3%** | 85 | **2.5%** | 13 | **4.8%** | 31 | **3.3%** | 84 | **3.6%** |
| | 55-59 | | 1,542 | **4.7%** | 1,220 | **4.7%** | 133 | **3.9%** | 16 | **5.9%** | 45 | **4.8%** | 128 | **5.5%** |
| | 60-64 | | 2,061 | **6.3%** | 1,648 | **6.4%** | 189 | **5.5%** | 17 | **6.2%** | 55 | **5.8%** | 152 | **6.5%** |
| | 65-69 | | 2,688 | **8.2%** | 2,112 | **8.2%** | 273 | **8.0%** | 20 | **7.3%** | 69 | **7.3%** | 214 | **9.2%** |

| | | | | | | | | | | | | | |
|---|---|---|---|---|---|---|---|---|---|---|---|---|---|
| | 70-74 | 3,815 | **11.6%** | 2,983 | **11.5%** | 382 | **11.1%** | 32 | **11.7%** | 122 | **12.9%** | 296 | **12.7%** |
| | 75-79 | 4,844 | **14.7%** | 3,878 | **15.0%** | 510 | **14.9%** | 30 | **11.0%** | 117 | **12.4%** | 309 | **13.3%** |
| | 80-84 | 5,211 | **15.9%** | 4,119 | **15.9%** | 568 | **16.6%** | 34 | **12.5%** | 166 | **17.5%** | 324 | **13.9%** |
| | 85-89 | 4,825 | **14.7%** | 3,791 | **14.7%** | 544 | **15.9%** | 37 | **13.6%** | 139 | **14.7%** | 314 | **13.5%** |
| | >=90 | 4,131 | **12.6%** | 3,320 | **12.8%** | 467 | **13.6%** | 32 | **11.7%** | 85 | **9.0%** | 227 | **9.7%** |
| **Gender** | Female | 16,450 | **50.1%** | 12,998 | **50.3%** | 1,651 | **48.1%** | 136 | **49.8%** | 500 | **52.8%** | 1,165 | **50.0%** |
| | Male | 16,258 | **49.5%** | 12,732 | **49.2%** | 1,778 | **51.8%** | 137 | **50.2%** | 447 | **47.2%** | 1,164 | **49.9%** |
| | Missing | 137 | **0.4%** | 132 | **0.5%** | 3 | **0.1%** | 0 | **0.0%** | 0 | **0.0%** | 2 | **0.1%** |
| **Ethnicity** | African | 221 | **0.7%** | 154 | **0.6%** | 29 | **0.8%** | 3 | **1.1%** | 7 | **0.7%** | 28 | **1.2%** |
| | Any other Asian background | 272 | **0.8%** | 189 | **0.7%** | 36 | **1.0%** | 3 | **1.1%** | 10 | **1.1%** | 34 | **1.5%** |
| | Any other Black background | 100 | **0.3%** | 67 | **0.3%** | 10 | **0.3%** | 5 | **1.8%** | 4 | **0.4%** | 14 | **0.6%** |
| | Any other White background | 1,576 | **4.8%** | 1,231 | **4.8%** | 160 | **4.7%** | 15 | **5.5%** | 53 | **5.6%** | 117 | **5.0%** |
| | Any other ethnic group | 308 | **0.9%** | 214 | **0.8%** | 35 | **1.0%** | 1 | **0.4%** | 9 | **1.0%** | 49 | **2.1%** |
| | Any other mixed background | 102 | **0.3%** | 73 | **0.3%** | 15 | **0.4%** | 1 | **0.4%** | 4 | **0.4%** | 9 | **0.4%** |
| | Bangladeshi or British Bangladeshi | 143 | **0.4%** | 109 | **0.4%** | 13 | **0.4%** | 0 | **0.0%** | 7 | **0.7%** | 14 | **0.6%** |
| | British, Mixed British | 26,771 | **81.5%** | 21,305 | **82.4%** | 2,771 | **80.7%** | 204 | **74.7%** | 749 | **79.1%** | 1,742 | **74.7%** |
| | Caribbean | 251 | **0.8%** | 161 | **0.6%** | 33 | **1.0%** | 4 | **1.5%** | 11 | **1.2%** | 42 | **1.8%** |
| | Chinese | 54 | **0.2%** | 37 | **0.1%** | 9 | **0.3%** | 0 | **0.0%** | 1 | **0.1%** | 7 | **0.3%** |
| | Indian or British Indian | 667 | **2.0%** | 464 | **1.8%** | 88 | **2.6%** | 7 | **2.6%** | 15 | **1.6%** | 93 | **4.0%** |
| | Irish | 376 | **1.1%** | 300 | **1.2%** | 36 | **1.0%** | 2 | **0.7%** | 11 | **1.2%** | 27 | **1.2%** |
| | Pakistani or British Pakistani | 473 | **1.4%** | 366 | **1.4%** | 39 | **1.1%** | 4 | **1.5%** | 21 | **2.2%** | 43 | **1.8%** |
| | White and Asian | 38 | **0.1%** | 26 | **0.1%** | 3 | **0.1%** | 0 | **0.0%** | 5 | **0.5%** | 4 | **0.2%** |
| | White and Black African | 25 | **0.1%** | 14 | **0.1%** | 7 | **0.2%** | 0 | **0.0%** | 1 | **0.1%** | 3 | **0.1%** |
| | White and Black Caribbean | 58 | **0.2%** | 40 | **0.2%** | 6 | **0.2%** | 1 | **0.4%** | 2 | **0.2%** | 9 | **0.4%** |
| | Missing | 1,410 | **4.3%** | 1,112 | **4.3%** | 142 | **4.1%** | 23 | **8.4%** | 37 | **3.9%** | 96 | **4.1%** |
| **NHS Region** | East of England | 2,955 | **9.0%** | 2,396 | **9.3%** | 309 | **9.0%** | 14 | **5.1%** | 68 | **7.2%** | 168 | **7.2%** |
| | London | 3,863 | **11.8%** | 2,810 | **10.9%** | 423 | **12.3%** | 31 | **11.4%** | 106 | **11.2%** | 493 | **21.1%** |
| | Midlands | 7,696 | **23.4%** | 6,059 | **23.4%** | 737 | **21.5%** | 64 | **23.4%** | 260 | **27.5%** | 576 | **24.7%** |
| | North East | 5,841 | **17.8%** | 4,535 | **17.5%** | 689 | **20.1%** | 43 | **15.8%** | 142 | **15.0%** | 432 | **18.5%** |

| | | | | | | | | | | | | | |
|---|---|---|---|---|---|---|---|---|---|---|---|---|---|
| | North West | 5,214 | **15.9%** | 4,257 | **16.5%** | 491 | **14.3%** | 48 | **17.6%** | 114 | **12.0%** | 304 | **13.0%** |
| | South East | 4,157 | **12.7%** | 3,366 | **13.0%** | 396 | **11.5%** | 46 | **16.8%** | 165 | **17.4%** | 184 | **7.9%** |
| | South West | 3,119 | **9.5%** | 2,439 | **9.4%** | 387 | **11.3%** | 27 | **9.9%** | 92 | **9.7%** | 174 | **7.5%** |
| | Missing | 0 | **0.0%** | 0 | **0.0%** | 0 | **0.0%** | 0 | **0.0%** | 0 | **0.0%** | 0 | **0.0%** |
| **IMD Quintiles** | 1 | 7,918 | **24.1%** | 6,236 | **24.1%** | 818 | **23.8%** | 63 | **23.1%** | 213 | **22.5%** | 588 | **25.2%** |
| | 2 | 6,923 | **21.1%** | 5,428 | **21.0%** | 728 | **21.2%** | 51 | **18.7%** | 198 | **20.9%** | 518 | **22.2%** |
| | 3 | 6,371 | **19.4%** | 5,049 | **19.5%** | 676 | **19.7%** | 55 | **20.1%** | 173 | **18.3%** | 418 | **17.9%** |
| | 4 | 6,232 | **19.0%** | 4,921 | **19.0%** | 645 | **18.8%** | 50 | **18.3%** | 191 | **20.2%** | 425 | **18.2%** |
| | 5 | 5,317 | **16.2%** | 4,168 | **16.1%** | 557 | **16.2%** | 53 | **19.4%** | 168 | **17.7%** | 371 | **15.9%** |
| | Missing | 84 | **0.3%** | 60 | **0.2%** | 8 | **0.2%** | 1 | **0.4%** | 4 | **0.4%** | 11 | **0.5%** |
| **Vaccine priority groups** | HSCW | 244 | **0.7%** | 172 | **0.7%** | 28 | **0.8%** | 4 | **1.5%** | 8 | **0.8%** | 32 | **1.4%** |
| | At risk*** | 5,396 | **16.4%** | 4,172 | **16.1%** | 540 | **15.7%** | 57 | **20.9%** | 178 | **18.8%** | 449 | **19.3%** |
| | Severely Immunosuppressed | 4,044 | **12.3%** | 3,108 | **12.0%** | 463 | **13.5%** | 37 | **13.6%** | 138 | **14.6%** | 298 | **12.8%** |
| | CEV | 15,813 | **48.1%** | 12,472 | **48.2%** | 1,719 | **50.1%** | 120 | **44.0%** | 407 | **43.0%** | 1,095 | **47.0%** |
| **Variant of most recent previous infection** | None | 27,270 | **83.0%** | 20,795 | **80.4%** | 3,212 | **93.6%** | 254 | **93.0%** | 870 | **91.9%** | 2,139 | **91.8%** |
| | Wild-type | 972 | **3.0%** | 872 | **3.4%** | 51 | **1.5%** | 2 | **0.7%** | 15 | **1.6%** | 32 | **1.4%** |
| | Alpha | 1,214 | **3.7%** | 1,086 | **4.2%** | 64 | **1.9%** | 5 | **1.8%** | 17 | **1.8%** | 42 | **1.8%** |
| | Delta | 1,186 | **3.6%** | 1,052 | **4.1%** | 53 | **1.5%** | 7 | **2.6%** | 21 | **2.2%** | 53 | **2.3%** |
| | Omicron | 2,203 | **6.7%** | 2,057 | **8.0%** | 52 | **1.5%** | 5 | **1.8%** | 24 | **2.5%** | 65 | **2.8%** |
| **Week Number** | 16 | 3,692 | **11.2%** | 2,638 | **10.2%** | 1,054 | **30.7%** | 0 | **0.0%** | 0 | **0.0%** | 0 | **0.0%** |
| | 17 | 3,337 | **10.2%** | 2,614 | **10.1%** | 718 | **20.9%** | 3 | **1.1%** | 2 | **0.2%** | 0 | **0.0%** |
| | 18 | 3,152 | **9.6%** | 2,593 | **10.0%** | 553 | **16.1%** | 4 | **1.5%** | 2 | **0.2%** | 0 | **0.0%** |
| | 19 | 3,002 | **9.1%** | 2,540 | **9.8%** | 449 | **13.1%** | 6 | **2.2%** | 7 | **0.7%** | 0 | **0.0%** |
| | 20 | 2,830 | **8.6%** | 2,471 | **9.6%** | 348 | **10.1%** | 4 | **1.5%** | 7 | **0.7%** | 0 | **0.0%** |
| | 21 | 2,332 | **7.1%** | 2,254 | **8.7%** | 55 | **1.6%** | 5 | **1.8%** | 18 | **1.9%** | 0 | **0.0%** |
| | 22 | 2,330 | **7.1%** | 2,206 | **8.5%** | 62 | **1.8%** | 23 | **8.4%** | 39 | **4.1%** | 0 | **0.0%** |
| | 23 | 2,320 | **7.1%** | 2,158 | **8.3%** | 57 | **1.7%** | 33 | **12.1%** | 72 | **7.6%** | 0 | **0.0%** |
| | 24 | 2,162 | **6.6%** | 1,910 | **7.4%** | 56 | **1.6%** | 60 | **22.0%** | 136 | **14.4%** | 0 | **0.0%** |

| | | | | | | | | | | | | |
|---|---|---|---|---|---|---|---|---|---|---|---|---|
| 25 | 2,501 | **7.6%** | 1,604 | **6.2%** | 42 | **1.2%** | 49 | **17.9%** | 241 | **25.4%** | 565 | **24.2%** |
| 26 | 2,075 | **6.3%** | 1,220 | **4.7%** | 24 | **0.7%** | 46 | **16.8%** | 163 | **17.2%** | 622 | **26.7%** |
| 27 | 1,804 | **5.5%** | 946 | **3.7%** | 12 | **0.3%** | 25 | **9.2%** | 156 | **16.5%** | 665 | **28.5%** |
| 28 | 1,308 | **4.0%** | 708 | **2.7%** | 2 | **0.1%** | 15 | **5.5%** | 104 | **11.0%** | 479 | **20.5%** |

*Dose 1 and 2 is recipients of ChAdOx1-S, BNT162b2 or mRNA-1273.

**Dose 3 or 4 is recipients of BNT162b2 or mRNA-1273 following any primary immunisation course.

***At risk is only those under 65

Percentages are column percentages. First row is shows row percentages.

Supplementary Table 4. Vaccine effectiveness estimates against hospitalisation for individuals aged 18 years and older in England (estimates from Figure 1a).

| Doses | Interval (weeks) | Controls | Cases | VE % (95% CI) | Cases | VE % (95% CI) | Cases | VE % (95% CI) |
|---|---|---|---|---|---|---|---|---|
| | | | BA.2 | | BA.4 | | BA.5 | |
| Unvaccinated | | 1,235 | 349 | -72.2 (-109.3 to -41.7) | 33 | -91.0 (-243.3 to -6.2) | 107 | -76.8 (-148.1 to -26.0) |
| Dose 1 | >0 | 406 | 73 | -21.9 (-65.8 to 10.4) | 4 | 30.1 (-112.3 to 77.0) | 24 | -10.5 (-86.0 to 34.3) |
| Dose 2 | 0-1 | 4 | 1 | n too small | 0 | n too small | 0 | n too small |
| | 2-14 | 35 | 3 | 53.0 (-62.5 to 86.4) | 1 | n too small | 1 | n too small |
| | 15-24 | 78 | 8 | 45.4 (-24.7 to 76.1) | 2 | n too small | 2 | n too small |
| | 25+ | 1,645 | 258 | Baseline | 20 | Baseline | 75 | Baseline |
| Final dose (Dose 3 or 4) | 0-1 | 1,443 | 163 | 51.7 (39.4 to 61.5) | 5 | 51.3 (-34.8 to 82.4) | 16 | 58.9 (26.7 to 77.0) |
| | 2-14 | 7,800 | 532 | 52.4 (43.2 to 60.1) | 61 | 56.8 (24.0 to 75.4) | 250 | 59.9 (45.6 to 70.5) |
| | 15-24 | 5,212 | 966 | 27.2 (14.4 to 38.1) | 29 | 22.2 (-42.8 to 57.6) | 85 | 38.7 (12.8 to 56.9) |
| | 25+ | 8,004 | 1079 | 9.8 (-6.0 to 23.3) | 118 | 1.5 (-63.1 to 40.5) | 387 | 23.3 (-1.5 to 42.1) |

Supplementary Table 5. Vaccine effectiveness estimates against hospitalisation for individuals aged 18 years and older in England (estimates from Figure 1b and 1c).

| Doses | Interval (weeks) | Controls | Cases | VE % (95% CI) | Cases | VE % (95% CI) |
|---|---|---|---|---|---|---|
| | | | | BA.2 | | BA.4 and BA.5 |
| Unvaccinated | | 1,235 | 349 | -72.1 (-109.1 to -41.6) | 409 | -69.9 (-113.7 to -35.1) |
| Dose 1 | >0 | 406 | 73 | -21.8 (-65.6 to 10.4) | 98 | -17.7 (-65.2 to 16.1) |
| Dose 2 | 0-1 | 4 | 1 | n too small | 0 | n too small |
| | 2-14 | 35 | 3 | 53.0 (-62.6 to 86.4) | 3 | n too small |
| | 15-24 | 78 | 8 | 45.4 (-24.7 to 76.1) | 11 | -17.2 (-169.2 to 49.0) |
| | 25+ | 1,645 | 258 | Baseline | 283 | Baseline |
| Final dose | 0-1 | 446 | 64 | 44.8 (24.8 to 59.6) | 15 | 38.9 (-16.0 to 67.8) |
| (Dose 3 or 4) | 2-14 | 3,959 | 335 | 50.3 (39.8 to 58.9) | 445 | 49.6 (37.5 to 59.4) |
| BNT162b2 | 15-24 | 4,223 | 805 | 26.5 (13.2 to 37.7) | 254 | 36.3 (19.1 to 49.8) |
| | 25+ | 7,375 | 1,052 | 9.5 (-6.5 to 23.1) | 1,319 | 16.7 (-0.3 to 30.7) |
| Final dose | 0-1 | 997 | 99 | 55.2 (41.8 to 65.6) | 44 | 55.4 (33.7 to 70.0) |
| (Dose 3 or 4) | 2-14 | 3,841 | 197 | 55.7 (45.1 to 64.2) | 470 | 62.2 (53.2 to 69.4) |
| mRNA-1273 | 15-24 | 989 | 161 | 30.3 (12.6 to 44.4) | 49 | 42.5 (14.3 to 61.4) |
| | 25+ | 629 | 27 | 23.5 (-17.5 to 50.2) | 151 | 42.0 (24.5 to 55.5) |
